# Large scale serum proteomics identifies proteins associated with performance decline and clinical milestones in Duchenne muscular dystrophy

**DOI:** 10.1101/2024.08.05.24311516

**Authors:** N.A. Ikelaar, A.M. Barnard, S.W.M. Eng, S. Hosseini Vajargah, K.C.H. Ha, H.E. Kan, K. Vandenborne, E.H. Niks, G.A. Walter, P. Spitali

## Abstract

Serum biomarkers are promising minimally invasive outcome measures in clinical studies in Duchenne muscular dystrophy (DMD). However, biomarkers strongly associated with clinical progression and predicting performance decline are lacking. In this study we aimed to identify serum biomarkers associated with clinical performance and able to predict clinical milestones in DMD. Towards this aim we present a retrospective multi-center cohort study including serum samples and clinical data collected in research participants with DMD as part of a natural history study at the University of Florida (UF) and real-world observations at Leiden University Medical Center (LUMC) between 2009-2022. The 7K SomaScan® assay was used to analyse protein levels in in individual serum samples. Serum biomarkers predicted age at loss of ambulation (LoA), age at loss of overhead reach (OHR) and age at loss of hand to mouth function (HTM). Secondary outcomes were the association of biomarkers with age, corticosteroid (CS) usage, and clinical performance based on the North Star Ambulatory Assessment (NSAA), 10 meter run velocity (10mrv), 6 minute walk (6MWT) and Performance of the Upper Limb (PUL2.0). A total of 716 serum samples were collected in 79 participants at UF and 74 at LUMC (mean[SD] age; 10.9[3.2] vs 8.4[3.4]). 244 serum proteins showed an association with CS usage in both cohorts independent of CS type and regimen, including MMP3 and IGLL1. 318 probes (corresponding to 294 proteins) showed significant associations with NSAA, 10mrv, 6MWT and/or PUL2.0 across both cohorts. The expression of 38 probes corresponding to 36 proteins such as RGMA, EHMT2, ART3, ANTXR2 and DLK1 was associated with risk of both lower and upper limb clinical milestones in both the LUMC and UF cohort. In conclusion, multiple biomarkers were associated with CS use, motor function and upper lower and upper limb disease milestones in DMD. These biomarkers were validated across two independent cohorts, increasing their likelihood of translation for use within the broader DMD population.

## Introduction

Duchenne muscular dystrophy (DMD) is the most common form of muscular dystrophy^1^. It is caused by variants in the *DMD* gene, resulting in an absence of functional dystrophin^2^. Symptoms begin in early childhood with delayed motor milestones and continue with progressive muscle weakness, scoliosis, reduced pulmonary function, and dilated cardiomyopathy. Untreated, most patients will lose ambulation by the age of 10 years^3,4^.

Current treatment consists of long-term corticosteroid (CS), initiated at age 4-5 years and recommended for life-long use^5^. Chronic use of CS has been shown to delay loss of ambulation, onset and progression of scoliosis, and, in combination with non-invasive ventilation, it increases life expectancy to 30-40 years of age^3,4^. Yet CS are not without negative metabolic effects^6^. Despite the huge progress in understanding the biology of the disease and the large investment in pre-clinical research, there is currently no cure for DMD. Multiple investigational drugs have failed to show significant improvement in participants’ performance in clinical trials, with only a few drugs receiving full and/or accelerated or conditional FDA approval such as microdystrophin gene therapy^7^, exon skipping^8^, vamorolone^9^ and givinostat^10^. While failures may be related to limited drug potency, it has also become clear that disease trajectories present high inter-individual variation, further reducing the power to detect a significant and clinically meaningful treatment effect on a single outcome measure in a typically short term, multi-center clinical trial. Several factors contribute to disease heterogeneity, including mutation type^11–14^, genetic modifiers^15^, and standards of care. Outcome measures have been shown to be affected by patient motivation^16^. These factors complicate accurate prediction of individual disease trajectories and thereby contribute to difficulty in clinical trial design. Therefore, being able to objectively predict long-term clinical outcomes based on short term evidence would greatly facilitate the evaluation of medicinal products.

Numerous serum biomarkers have been proposed for DMD^17–19^, however the focus has mostly been in the identification of cross-sectional differences between unaffected controls and individuals with DMD. Monitoring biomarkers, evaluated in series, are particularly lacking. Research efforts focused on the analysis of muscle damage biomarkers^20–22^, and in monitoring response to (micro-)dystrophin restoration therapies^23,24^. However, serum biomarkers with prognostic value are currently lacking.

The challenge and paradox relies on a disconnect between the early window of opportunity to treat patients (roughly 3-8 years of age, in which most of the muscle mass is available for therapeutic applications aimed at prevention vs reversal), the observable decline in patients function (typically starting around 8-9 years of age), and the observation of disease milestones such as loss of ambulation (between 10-16 years of age). Initial attempts to determine longitudinal trajectories in serum biomarkers related to disease progression^22,25,26^ have been limited by the small number of individuals, lack of follow-up samples, and incomplete collection of clinical data.

Therefore, discovery studies in large and well-characterized cohorts are needed to identify associations between biomarker signatures and clinical performance and to define the context of use for the biomarker signature. Importantly, prognostic biomarkers that can predict disease milestones over longer time frames could enrich clinical trial design. Patients recruitment can be tailored to individuals who are more likely to experience the milestone during the study. This is particularly relevant as participants are usually enrolled in clinical trials before the age of 8, in early stages of disease progression, but when little decline can be normally observed within the duration of an interventional study of 12 months. Connecting short term changes in biomarker signatures to the likelihood of long-term decline would help bridge this gap. Therefore, in this study we aimed to identify serum biomarkers associated with clinical outcomes and their prognostic value for clinically relevant milestones for both lower and upper limb function.

## Methods

### Study Cohort, design and outcomes

This was a retrospective, multi-center, cohort study including serum samples and clinical data collected from individuals with DMD participating in research protocols at the University of Florida (UF) and at the Leiden University Medical Center (LUMC) between 2009-2022. We included 407 serum samples from 74 individuals with DMD aged 4 – 24 years at LUMC and 295 serum samples from 79 individuals with DMD aged 5 – 22 years at UF. At LUMC, samples were collected during yearly visits to the outpatient clinics as part of the standard of care. At UF, samples were collected as part of an optional biosample addition to research visits conducted for the ImagingDMD natural history study (NCT01484678). Written consent was obtained from all participants or their caregivers as described in protocol B22.013 at LUMC and protocols IRB201500981 and IRB201700056 at UF, which were approved by the respective regulatory boards at both sites.

Clinical data were obtained at the same clinic or research visit as serum sample collection. Data included age at sample collection, CS use at the time of sample collection, and performance on tests of function. CS information was categorized by use (treated or untreated), type (deflazacort, prednisone, or other), regimen (daily or intermittent – defined as 10 days on/10 days off or weekend dosing), and dose. Motor function tests included the North Star Ambulatory Assessment (NSAA), 10 meter run/walk velocity (10MRV), 6 minute walk test (6MWT), and Performance of the Upper Limb (PUL2.0)^27–30^. Three disease milestones were recorded: age at loss of ambulation (LoA), age at loss of overhead reach (OHR), and age at loss of hand to mouth (HTM). LoA was defined similarly in both cohorts with LoA based on patient reported inability to walk 5 meters unaided at home at LUMC and inability to traverse 10 meters unaided within 45 seconds at UF. OHR and HTM were primarily derived from PUL2.0 scores and occasionally from patient reported data.

### Samples collection and proteomic analysis

Serum samples were collected and prepared according to standard phlebotomy procedures. Samples at LUMC were left to clot for ∼30 minutes, followed by 10 min centrifugation at 2350g, while clotting time was 30 minutes at UF, followed by centrifugation at 3000 rpm for 15 min. At the LUMC, sample aliquots were frozen at –20°C for 1-2-months and then transferred to –80°C for long term storage, while at UF, samples were immediately stored at –80°C. Serum samples were analysed by the SomaScan® proteomic platform at SomaLogic (Boulder, Colorado, USA)^31^. SomaScan uses aptamers as affinity reagents to detect and quantify proteins in complex mixtures such as serum. Aptamers are nucleic acids that are able to bind specific protein targets with high specificity and affinity. The pipeline is built to keep targets engaging aptamers, while washing away the unbound ones. Detection of the aptamers is then performed using an array with probes complementary to the aptamer sequence. At the time of this research, this platform included 7596 aptamers capable of detecting 6628 proteins. As part of the assay, extensive quality control metrics are put in place including internal and external controls. A total of 9 samples (2 LUMC / 7 UF) did not pass SomaLogic’s internal quality control and were excluded from further analysis, results, and data tables.

### Statistical analysis

Linear mixed effects models (LMEM) were constructed for each site and probe to predict that probe’s expression given age and CS use as follows:

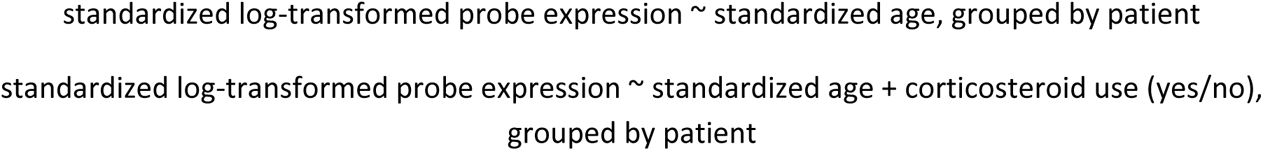

LMEMs were then used to predict motor function tests, controlling for age and CS use as follows:

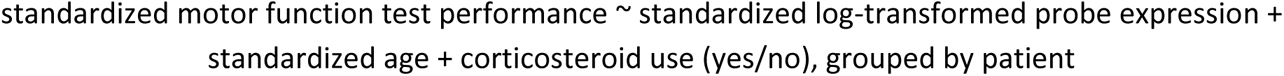

LMEMs were conducted using the *pymer4* package, version 0.7.8, a Python interface for the *lme4* R package, version 1.1-31^32,33^. To determine how generalizable the significant associations between probes and motor function tests were, for each site, the associated LMEMs were used to predict values of the motor function tests for the other site. That is, for LUMC models, UF predicted values were generated using UF data, and for UF models, LUMC predicted values were generated using LUMC data. Predicted values were compared against original values via reconstruction accuracy:

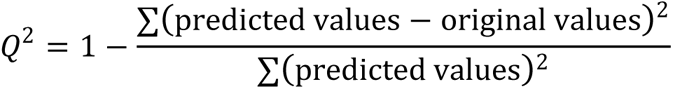

Significant probes whose validation Q^2^ values were within the top 5% of Q^2^ values (i.e., at or above the 95^th^ percentile) were kept for prediction of the clinical milestones (age at LoA, loss of OHR, and loss of HTM). These probes were grouped based on the milestone being predicted for each site. For LoA, probes associated with NSAA, 10MRV, and/or 6MWT were considered. For loss of HTM and OHR, probes associated with PUL2.0 were considered. For each site, clinical milestone, and candidate probe, a Cox proportional hazards model was constructed to predict that milestone using the following specification:

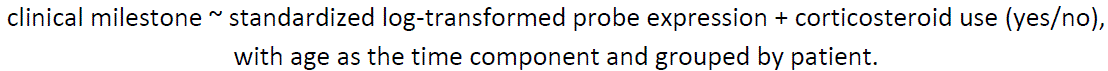

Modelling was conducted using the *lifelines* Python package, version 0.27^34^. All p-values underwent false discovery rates (FDRs) adjustments to account for multiple hypothesis testing. Probes with FDRs < 0.05 were considered to be significantly associated with the respective outcomes. A probe was significantly associated with an outcome if its FDR was <0.05.

## Data availability

Anonymized data can be made available to qualified investigators on request. Requests should be in line with the approved ethical protocol.

## Results

After retaining samples passing quality control, a total of 153 males with DMD and 693serum samples were included in the study (Table 1). LUMC participants were significantly younger at the first sample visit compared to UF participants (mean[SD]; 8.4[3.4] vs 10.9[3.2], p<0.001). An average of 4.3 samples per participant were analysed, with longer follow-up duration in the LUMC cohort (5.7[3.6] vs 3.4[2.5] years, p<0.001). An intermittent (10 days on/10 days off of pred (or Deflazacort) CS regimen was most common at LUMC while daily dosing was most common at UF (prednisone and deflazacort); however, age at start of CS treatment was comparable across sites. Only a few patients remained CS naïve for the entire study period (4.1% vs 3.2%). The percentage of patients meeting one of the 3 clinical milestones at first sample was higher at LUMC compared to UF (LoA: 18.9% vs 9.6%, loss of OHR: 16.0% vs 12.2%, loss of HTM: 5.3% vs 0.0%). For the LUMC cohort, 6660 probes passed quality control compared to 6690 for the UF cohort.

**Table 1.** Cohort characteristics.

|  | LUMC<br>n = 74 | UF<br>n = 79 |
| --- | --- | --- |
| *Samples, n* | 405 | 288 |
| *Age at first sample, years, mean (SD)* | 8.4 (3.4) | 10.9 (3.2) |
| *Follow-up duration, years, mean (SD)* | 5.7 (3.6) | 3.4 (2.5) |
| *Samples per patient, n, mean (SD)* | 5.5 (2.9) | 3.6 (2.0) |
| *Age at start of corticosteroids, years, mean (SD)* | 5.6 (1.6) | 5.4 (1.7) |
| *Corticosteroid treatment at first sample, n (%)* |  |  |
| • Not recorded | 1 (1.4%) | 3 (3.8%) |
| • Daily | 2 (2.7%) | 66 (83.5%) |
| • Intermittent | 49 (66.2%) | 4 (5.1%) |
| • None | 22 (29.7%) | 6 (7.6%) |
| *Dystrophin variants, n (%)* |  |  |
| • Exonic deletion | 49 (66.2%) | 49 (62.0%) |
| • Exonic duplication | 10 (13.5%) | 8 (10.1%) |
| • Nonsense variant | 13 (17.6%) | 12 (15.2%) |
| • Splice site variant | 2 (2.7%) | 3 (3.8%) |
| • Other small point variants | — | 5 (6.3%) |
| • Variant unknown | — | 2 (2.5%) |
| *Weight at first sample, kg, mean (SD)* | 32.6 (18.6) | 35.1 (12.2) |
| *Height at first sample, cm, mean (SD)* | 127.2 (19.2) | 126.9 (10.7) |
| *BMI at first sample, mean (SD)* | 18.3 (4.4) | 21.2 (5.4) |
| *Ambulant patients at first sample, n (%)* | 59 (79.7%) | 68/78 (87.2%)<br>(1 unknown) |
| *Able to perform overhead reach at first sample, n (%)* | 64 (86.5%) | 63/70 (90.0%)<br>(9 unknown) |
| *Able to perform hand to mouth at first sample, n (%)* | 73 (98.6%) | 74/75 (98.7%)<br>(4 unknown) |

### Serum protein associations with age and corticosteroid usage

Given the progressive nature of DMD and the availability of longitudinal visits, we first identified proteins associated with age. 4,796 probes (4436 proteins) were significantly associated with age in the LUMC cohort and 2,668 probes (2498 proteins) were significantly associated with age in the UF cohort (FDR<0.05)(Fig. 1A-B); with overlap of 2,317 probes (2186 proteins) between both cohorts (Fig. 1C). 2,251 probes showed concordant directional change between cohorts, with the coefficients generally being higher in the LUMC cohort (Fig. 1D). Multiple muscle proteins were negatively associated with age including, but not limited to, CK-MM, MYOM3, MYOM2, TRIM72, MYL3, BIN1, TTN, LAMA2, TPM3, ACTN2, DES, CA3, MYL3 and TNNI2 (Fig1E-H). Markers of fibroadipogenic progenitors (PDGFRA), fibrosis and extracellular matrix (collagens such as COL1A1, COL2A1, COL3A1, COL6A1, COL6A2, COL6A3, TIMP1), several BMPs (BMP-1,4,5,6,7) and protein synthesis pathways (AKT1, AKT2, IGF2), also decreased with increasing age.

**Figure 1.**
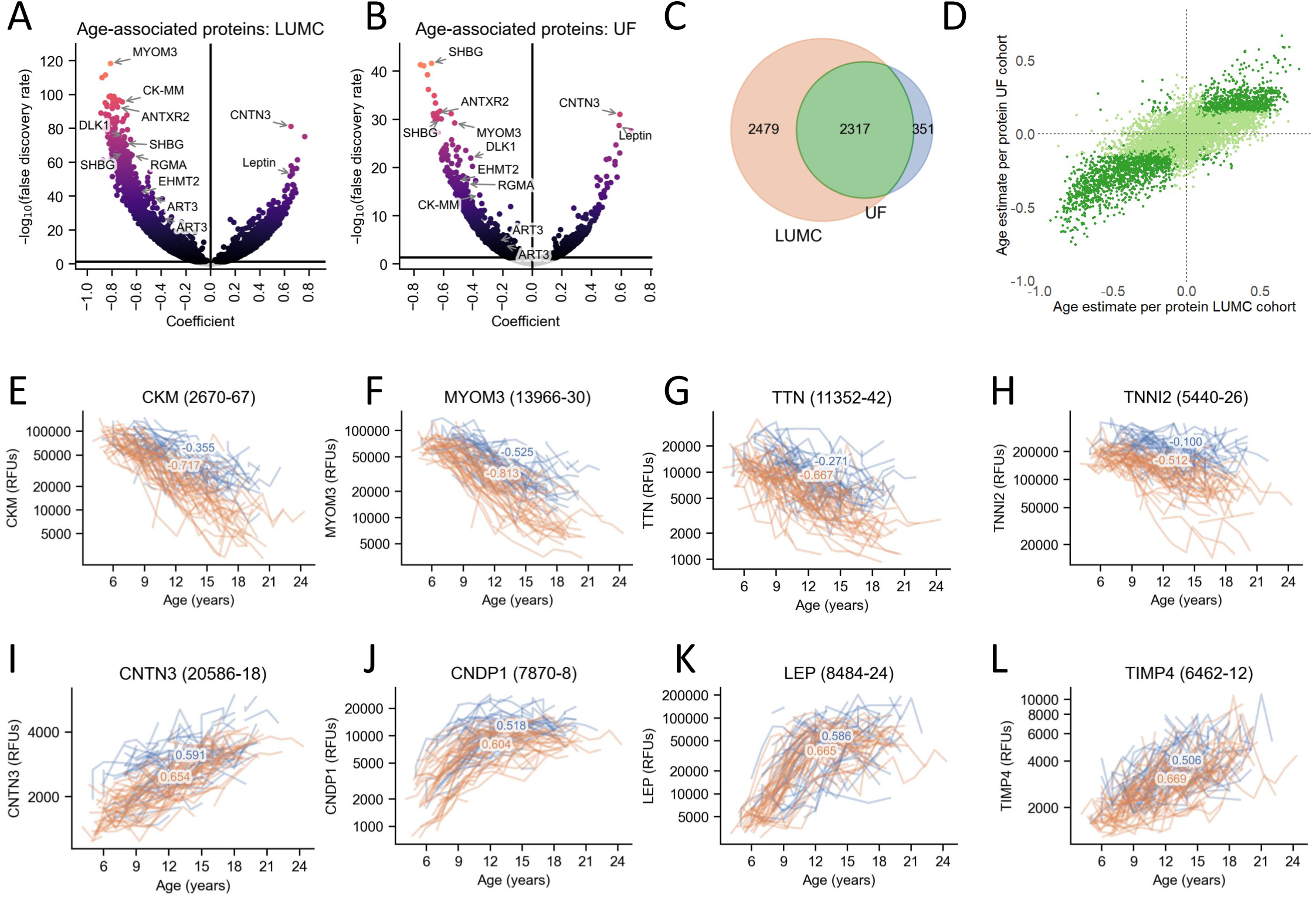
Associations of Serum Proteins with Age in Individuals with DMD. Volcano plots demonstrate the proteins from DMD serum that are associated with age in A) the LUMC cohort, and B) the UF cohort. C) Venn diagram showing the number of proteins associated with age in common between the LUMC and UF cohorts, with D) a plot of coefficients, indicating the direction of protein association with age, in the LUMC versus UF cohorts. Plots E-H highlight the longitudinal trajectories of protein levels over time in select proteins that decrease with increasing age such as CKM, MYOM3, TTN and TNNI2, while plots I-L show trajectories of select proteins that increase with increasing age such as CNTN3, CNDP1, LEP and TIMP4. Each trajectory plot includes the gene symbol and aptamer identification number. DMD, Duchenne muscular dystrophy; LUMC, Leiden University Medical Center; UF, University of Florida.

Among the proteins with expression that significantly increased with age, we identified proteins present in the central nervous system such as CNTN3 and CNDP1, proteins expressed in adipose tissue (LEP, GHR, FABP4, ADIPOQ, CNTFR, CFD and TIMP4, PNLIPRP1, PNLIPRP2, PNLIP), and several cytokines (CCL14, IFNA6, BMPR2, CXCL8, CCL3L1, IL26, IL5RA, CCL1, CCL16, CCL15, IL10RA, IL31RA, IL17RE) (Fig1I-L). Two different probes for IGF1 had had opposing associations with age, and several IGF binding proteins showed discordant trajectories, with IGFBP1 and IGFBP2 decreasing with age andIGFBP5 and IGFBP6 increasing with age. Coefficients of all significant, concordant proteins associated with age are provided in Supplementary Table 1.

Given that the majority of participants had chronic exposure to CS, we sought to identify a protein expression signature related to exposure to CS while correcting for age. 846 probes (790 proteins) in the LUMC cohort and 396 probes (364 proteins) in the UF cohort were significantly associated with CS use (FDR < 0.05)(Fig. 2A-B). Furthermore, 244 probes (227 proteins) were shared across sites (Fig. 2C). The number of proteins associated with steroid use in the UF cohort was smaller due to the small number of patients untreated (n=5) in this cohort compared to the untreated group in the LUMC cohort (n=22), suggesting that the number of proteins identified as significantly associated with the treatment is not a measure of the treatment effect. This is confirmed by the comparison of the coefficients shown in Fig.2D, showing larger effects for the UF cohort in line with the higher dosage (mostly daily) compared to the LUMC cohort (mostly intermittent). Among the proteins we identified (CD23) and (MMP3)^6^ previously observed in DMD, as well as newly identified proteins such as IGLL1 immunoglobulin lambda-like polypeptide 1 (IGLL1) and repulsive guidance molecule A (RGMA)(Fig. 2E-G). To identify whether the identified associations were potentially related to efficacy or safety effects, we assessed the directionality of the signature for age and steroids. Proteins showing discordant association (such as declining with age as disease progresses and increased by CS treatment) were considered to relate to the steroid efficacy signature, while concordant effects were considered to belong to the safety signature (Fig.2I). A total of 44 proteins showed discordant age and CS effects (potential efficacy signature) across the 2 cohorts (Supplementary table 2); this list included previously identified proteins such as ANGPT2 (2 independent aptamers), proteins related to lipoproteins transportation (APOA4, APOC3, APOE) as well as newly identified recognized by 2 independent aptamers such as DLD, ART3, NDUFA2, OLFM2 as well as 2 members of the RGM family (RGMA and RGMC)(Fig.2H). A total of 172 proteins showed concordant sign for age and CS effects in both LUMC and UF cohorts (potential safety biomarkers; Supplementary Table 3). This list included previously identified MMP3, Afamin, IGFBP-5, proteins involved in SMAD signalling as well as new proteins belonging with IGF binding properties and metalloproteinases.

**Figure 2.**
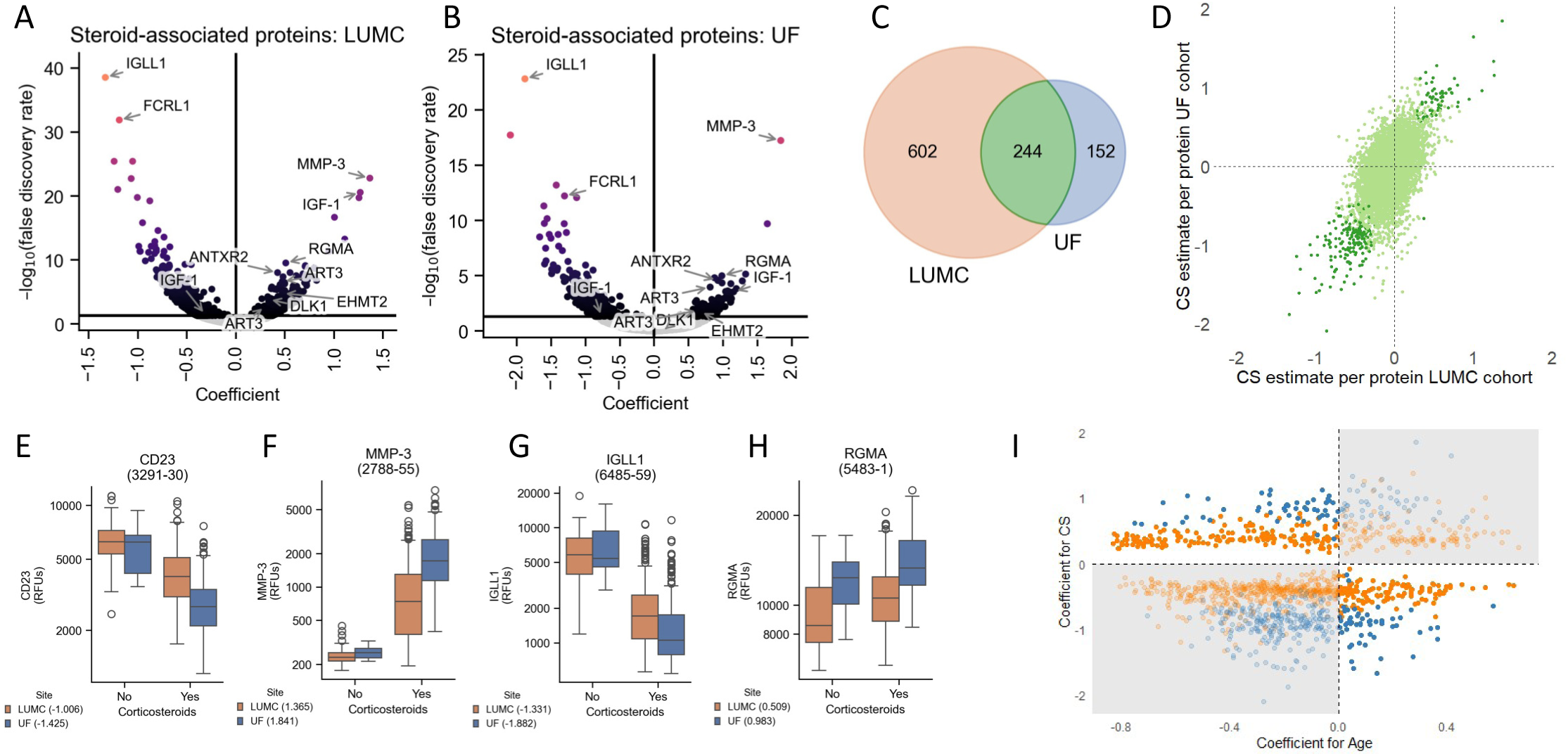
Associations of Serum Proteins with Corticosteroid use in Individuals with DMD. Volcano plots demonstrate the proteins from DMD serum that are associated with corticosteroid use in A) the LUMC cohort, and B) the UF cohort. C) Venn diagram showing the number of proteins associated with corticosteroid use in common between the LUMC and UF cohorts, with D) a plot of coefficients, indicating the direction of protein association with corticosteroid use, in the LUMC versus UF cohorts. Plots E-H show differences in protein levels between serum from CS treated versus untreated individuals with DMD in two previously identified proteins and two proteins identified in this study. I) Scatterplot showing the relationship between age and steroid treatment coefficients for proteins significantly associated with CS treatment. Proteins showing discordant coefficients for age and CS treatments were considered as efficacy biomarkers, while with discordant coefficients were considered as safety biomarkers (grey shaded). Orange dots represent estimates for the LUMC cohort, while blue dots represent estimates for the UF cohort. CS, corticosteroid; DMD, Duchenne muscular dystrophy; LUMC, Leiden University Medical Center; UF, University of Florida.

### Serum protein associations with motor performance

Longitudinal trajectories for the NSAA, 10MRW, 6MWT, and PUL 2.0 differed between the two sites, with a shift towards earlier decline in motor performance in the LUMC cohort (Fig 3A-D). To assess which proteins were associated with clinical severity, we evaluated associations with each functional test in each cohort. 2,005 probes (1882 proteins) showed a significant association with at least one motor function test in the LUMC cohort compared to 483 probes (454 proteins) in the UF cohort (FDR < 0.05). 318 probes (294 proteins) were shared across the two sites, with the majority of significant associations found for lower limb scales, consistent with the higher number of observations and the larger degree of functional decline experienced in the cohorts. Cross-correlation of the coefficients showed concordant directionality between the 2 cohorts, especially for the lower limb scales (Fig. 3E).

**Figure 3.**
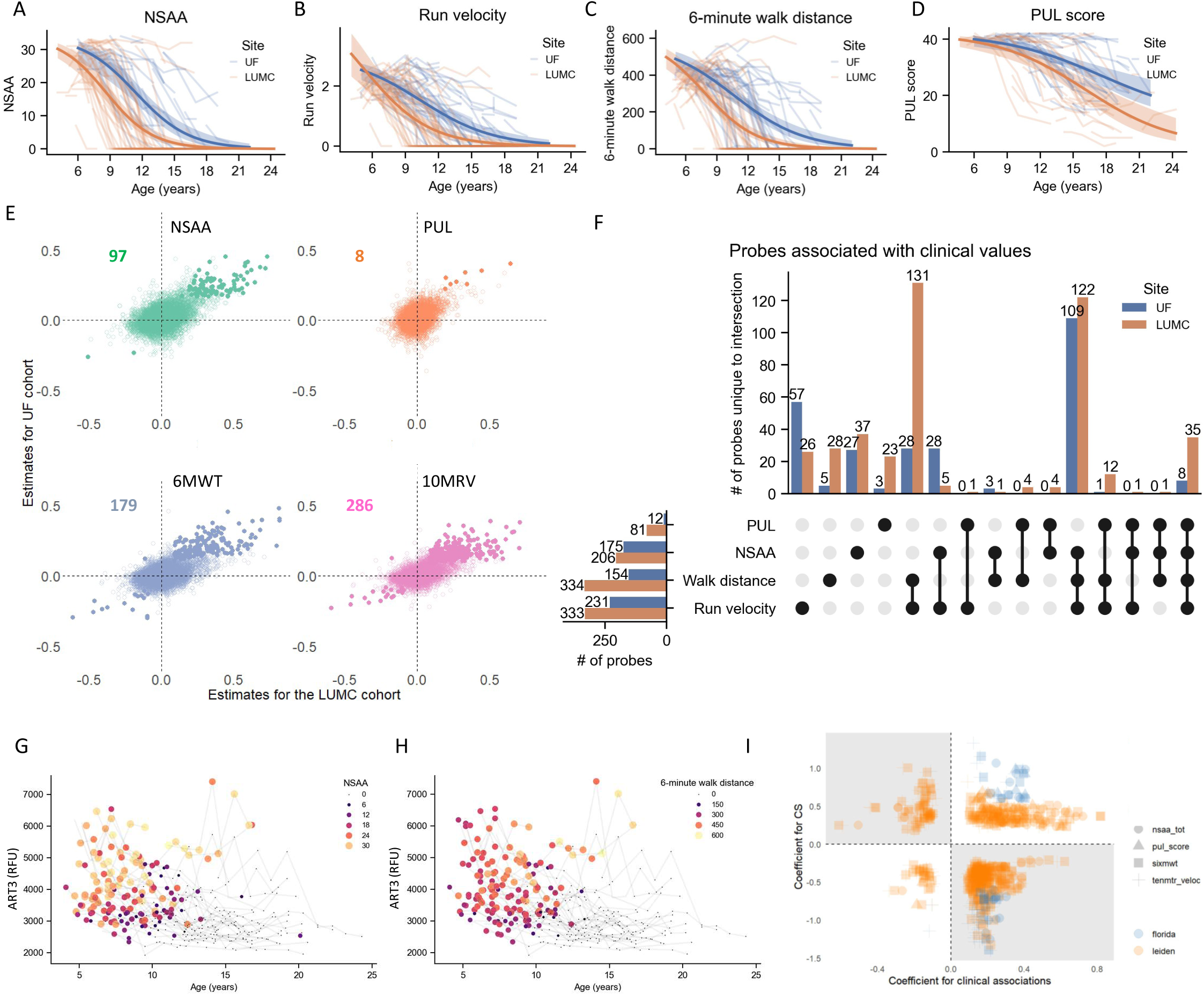
Associations of Serum Proteins with Performance on Tests of Motor Function in Individuals with DMD. A-D) Longitudinal trajectory plots of NSAA results, 10MRV, 6MWD, and PUL 2.0 score from both cohorts. E) Plots of coefficients, indicating the direction of protein association with motor function test performance in the LUMC versus UF cohorts. Highlighted data points represent significant associations. The number of significant associations is mentioned for each panel. F) UpSet plot demonstrating the intersection of significantly associated probes with each of the four tests of motor function. G-H) Trajectory plots showing the relationship between ART3 and either NSAA (G) or 6MWT (H) over the age in the LUMC cohort. I) Scatterplot showing the relationship between the coefficient estimates for proteins significantly associated with steroid treatment and performance test. Proteins showing concordant coefficients for performance tests and CS treatments were considered as efficacy biomarkers (e.g. a protein associated with both high functional scores and further elevated by steroid treatment), while with concordant coefficients were considered as safety biomarkers (grey shaded). Orange dots represent estimates for the LUMC cohort, while blue dots represent estimates for the UF cohort. Shapes indicate the association for each performance test. 6MWD, 6-minute walk distance; 10MRV, 10-meter run velocity; CS, corticosteroid; DMD, Duchenne muscular dystrophy; LUMC, Leiden University Medical Center; NSAA, North Star Ambulatory Assessment; PUL, Performance of the Upper Limb; UF, University of Florida.

We then assessed how generalizable models trained in each cohort were by using the other cohort for validation, selecting the 5% most performant proteins for each scale. Validation reconstruction accuracies were higher for lower limb function with Q^2^ values comparable across both cohorts for the 10MRV, while accuracies were higher for models trained on the LUMC cohort for the 6MWT and higher for UF for the NSAA. 122 probes (116 proteins) in the LUMC cohort and 109 probes (105 proteins) in the UF cohort significantly and accurately predicted all 3 scales used to monitor lower limb function (Fig. 3F). Supplementary Table 4 provides a list of proteins with significant associations across scales and cohorts. ART3, RGMA and DLD showed highly significant associations and accurate predictions across multiple motor function tests (Fig. 3G-H). To refine the efficacy and safety signature identified in the steroid analysis we assessed whether cross-correlation between steroid and clinical association coefficients; we identified 919 pairs where a protein was significantly associated with a clinical score and with steroid treatment across both cohorts. Concordant coefficients are in this case expected to be considered efficacy biomarkers, while discordant coefficients are expected to be adverse effects (Fig. 3I). A chi-squared test showed that concordant signs were significantly enriched in the previously defined efficacy signature, while discordant signs were enriched in the safety signature (P<10^-15^), with only 835 of the 919 pairs being properly assigned.

### Prediction of clinical milestones

Given the association with continuous clinical scores, we sought to determine whether the identified proteins had prognostic value for clinically meaningful disease milestones such as age at LoA, loss of OHR, and loss of HTM. All clinical milestones were reached at significantly earlier ages and in a greater percentage of patients in the LUMC cohort than in the UF cohort (Table 1, Fig. 4A-B). We identified 41 probes / milestone associations. Among these, 3 of the probes targeting RGMA, ENPP5 and RGS21 were associated with 2 different milestones (FDR <0.05). RGMA was associated with lower and upper limb milestones across the 2 cohorts, while RGS21 (UF) and ENPP5 (LUMC) were cohort-specific. Associations for DLK1 and ART3 were confirmed by 2 independent probes: DLK1 was found to be associated with OHR in the UF cohort and ART3 with LOA in the LUMC cohort (Fig.4C). The direction of log Hazard Ratios (logHR) were conserved for these proteins across probes and outcomes. Notably, RGMA, ANTXR2, EIF4G1, CAMK2A had large negative logHRs and PTPRD, NCAM had large positive logHRs. Kaplan-Meier plots for RGMA are presented across all 3 milestones for both LUMC and UF cohorts (Fig 4D). Supplementary Table 5 lists all proteins significantly predicting lower and upper limb milestones. Since treatment with steroids is known to delay these milestones, we assessed whether logHR and steroid effects show discordant effects. Figure 4E illustrates how estimates are indeed anti-correlated for all but 2 proteins, supporting the use of these biomarkers to evaluate the risk of meeting these milestones.

**Figure 4.**
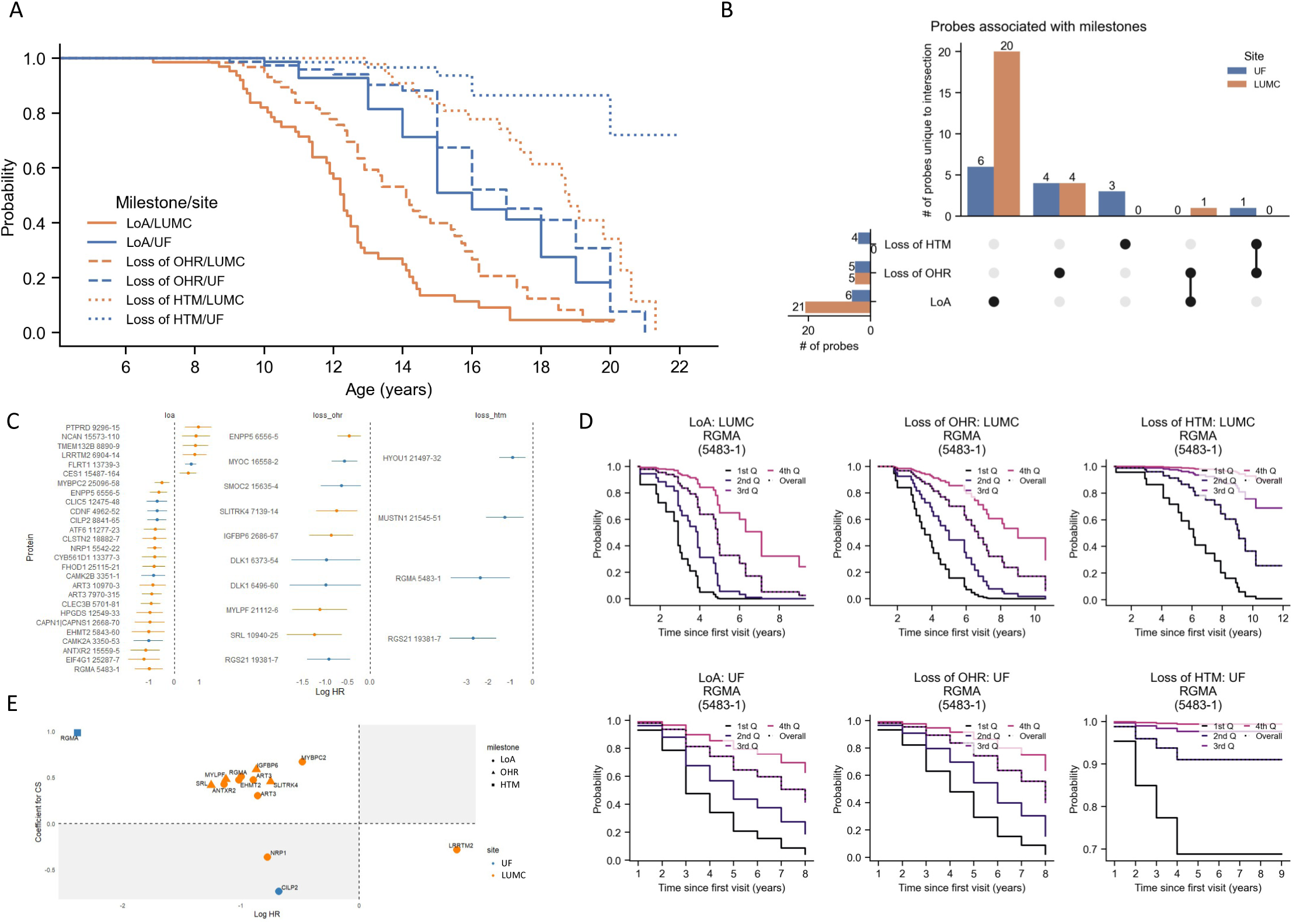
Associations of Serum Proteins with Clinical Milestones. A) Kaplan Meier curves showing the probability of LoA, loss of OHR, and loss of HTM by age in each cohort. B) UpSet plot of the intersection of significant probes with each clinical milestone. C) Log hazard ratios of the protein probes significantly associated with at least 1 clinical milestone. D) Kaplan Meier curves for loss of each clinical milestone for RGMA by years since first sample collection. E) Correlations between protein probe log hazard ratios and use of corticosteroids. HTM, hand-to-mouth; LoA, loss of ambulation; OHR, overhead reach.

Among the multiple proteins associated with functional tests and disease milestones, we shortlisted a few based on the consistency and strength of the associations across cohorts (Table 2). Proteins ANTXR2, ART3, EHMT2 and RGMA showed a % risk increase for disease milestones between 136% and 981% for a decrease of one unit (in log scale). Treatment with steroids corresponded to an increase in the biomarker level corresponding to a reduced the risk % up to 90% with highest normalization of risk for the upper limb HTM milestone; normalization for the risk % of LoA was in all cases estimated between 22% and 39%. We further estimated how monitoring proteins in blood can help evaluate the yearly increase in risk % for the milestones and observed yearly risk increase between 43% and 215%.

**Table 2.** Shortlisted proteins with consistent association across performance tests, milestones and cohorts. The coefficient estimates for the indicated milestone are obtained from the time to event analysis, while the coefficient for CS and age were obtained by the linear mixed effect models including both CS and age. We further show the risk % for the indicate milestone for a unit decrease in the protein (in log scale) and how the risk is reduced by treatment with CS and increased per year.

| Gene Symbol | Probe | Site | Milestone | Log HR | $\beta$ CS | $\beta$ age | Risk % increase with 1 log decrease | Risk % reduction with CS treat | Yearly risk % increase |
| --- | --- | --- | --- | --- | --- | --- | --- | --- | --- |
| ANTXR2 | 15559-5 | LUMC | LoA | -1.14 | 0.43 | -0.76 | 213.54 | 38.50 | 138.38 |
| ART3 | 7970-315 | LUMC | LoA | -0.89 | 0.47 | -0.41 | 144.55 | 34.49 | 43.83 |
| ART3 | 10970-3 | LUMC | LoA | -0.86 | 0.30 | -0.48 | 135.79 | 22.67 | 51.19 |
| EHMT2 | 5843-60 | LUMC | LoA | -1.02 | 0.47 | -0.60 | 176.06 | 38.11 | 83.08 |
| RGMA | 5483-1 | UF | HTM | -2.38 | 0.98 | -0.48 | 981.79 | 90.38 | 215.03 |
| RGMA | 5483-1 | LUMC | LoA | -1.00 | 0.51 | -0.64 | 170.79 | 39.78 | 89.96 |

## Discussion

Large-scale serum biomarker discovery is now possible in DMD with the availability of high-throughput proteomics platforms. In this retrospective study, we had the unique opportunity to pair robust and comprehensive clinical data from two large, independent international cohorts of individuals with DMD to serum levels of 6,628 proteins. The availability of extensive longitudinal serum samples and clinical data not only allowed for analysis of protein signatures associated with age and corticosteroid usage, but also allowed for identification of novel proteins predictive of clinical function and clinical milestones. Proteins like RGMA were predictive of both lower and upper limb clinical milestones such as LoA and loss of HTM. Furthermore, several proteins were associated with increased risk of single clinical milestones such as ART3 for LoA. The ability to compare these results across the two independent cohorts enabled for cross-validation of our findings.

Prior serum biomarker discovery in dystrophinopathies has largely been limited to smaller cohorts, targeted protein selection for analysis, and limited longitudinal data. These studies have been crucial for building our understanding of the disease’s pathogenic processes. Key differences between individuals with and without DMD have been reported, including several proteins associated with age, shedding light on the ongoing pathogenic processes. It is well-known with DMD that serum CK (activity and protein levels) decrease with age, reflecting muscle damage and loss of muscle mass. Additional proteins decreasing with age have subsequently been identified including MDH2, MYL3, CA3, MYOM3, COL1A1, TTN, TNNT2, ETFA, TNNT3, and MAP4^26,35^. Proteins that have increasing expression with age have included C4A, C4BPA, and GSN, among others^35^. A small number of studies have also utilized the SomaScan® platform to perform large-scale protein biomarker analysis in DMD, identifying additional proteins associated with age/time such as LEP, CFTNR, FABP3, and TNNI2^25,36^. Complementing and largely validating the findings of these studies, we identified previously described proteins such as CK and MYOM-3 showing steep decline in protein levels, but also substitution of muscle with fibro-adipogenic tissue with markers such as Collagen 1 and 2 declining along with markers of fibro-adipogenic progenitors such as PDGFRA^37^. We also observed adipogenic markers such as LEP, GHR and ADIPOQ with increased expression along with a complement and inflammation signature.

The CS signature encompassed steroid engagement biomarkers such as MMP3 and IGLL1, which showed the strongest association with CS usage, both in the daily and the intermittent regimen. Previous research has also identified elevated MMP3 in patients with DMD treated with CS, while IGLL1 has not yet been described in relation to DMD^38^. However, it has very recently been shown that weekend CS use in adults with limb girdle muscular dystrophy reduces IGLL1 in the presence of MMP3 elevation^39^. Given the role of IGLL1 in B cells, the reduction observed in combination with the CS treatment could signal the immunosuppression expected in these patients due to the drug.

Treatment with corticosteroids contributed to normalize as well as exacerbate the disease progression signature. An exacerbation of the signature related to age was observed for previously reported safety biomarkers such as MMP3, Afamin, and IGFBP5^6^, and for certain apolipoproteins such as APOA2, APOL1, APOA5. A normalization was observed for other apolipoproteins such as APOE4, APOC3 and APOE, suggesting that steroid treatment could directly normalize the dyslipidemia with effects on APOE4, APOC3, and APOE, while exacerbation of the APOA2, APOL1, and APOA5 signature could highlight how steroids affect lipid metabolism and potentially cardiovascular health. The compensatory effect of APOE is also underscored by the more severe phenotype observed in *mdx* ApoE double knock-out mice^40,41^. We also found that other previously reported proteins such as ANGPT2^6^, and proteins (RGMA, DLK1, ANTXR2, and ART3) that were decreased with disease progression in both UF and Leiden cohorts were higher in the CS groups compared to untreated.

A major strength of this study was the ability to identify serum biomarkers predictive of clinical function. RGMA and ART3 were all directly related to muscle performance as measured by both upper (PUL2.0) and lower limb (NSAA, 6MWT, 10MRV) functional outcomes. RGMA, ANTXR2, EHMT2 and ART3 were found to have large negative logHR when considering the loss of certain clinical milestones. To further illustrate this, we stratified the population according to RGMA levels and found that the decreasing levels of RGMA clearly shifted the Kaplan-Meier curve towards earlier LoA and OHR.

RGMA is part of the repulsive guidance molecule family of glycoprotein-1 (GP1) anchor proteins that is mostly expressed in the central nervous system and muscle tissue according to the human protein atlas gene expression data. Originally described as playing an important role in neurogenesis by guiding axonal growth and as an important target of neuronal survival, it has now also been identified to play a role in myogenesis^42,43^. RGMA has been proposed to play a central role in regulating cellular hypertrophy and hyperplasia^44^. RGMA has previously been found to have an association with Spinal and bulbar muscular atrophy (SBMA)^45^, Parkinson disease, Alzheimer disease, multiple sclerosis, and cerebrovascular accidents, as well as an association with upper limb function measured by elbow flection in DMD^25,42^.

ANTXR2, also known as capillary morphogenesis protein 2 (CMG2) plays an important role in cellular interaction by binding collagen IV and laminin, suggesting involvement in extracellular matrix adhesion. It is expressed in multiple tissues, including muscles. Loss of function variants in ANTXR2 cause hyaline fibromatosis syndrome and ANTXR2 knockout mice show collagen VI accumulation in the uterus^46^. This could implicate ANTXR2 as playing a role as a collagen VI regulator, and possible involvement in muscle homeostasis.

ART3, known as ADP-Ribosyltransferase 3, is mainly expressed in skeletal muscle tissue according to GTEx and human protein atlas databases. Gene expression data obtained from human individuals as well as in Chinese Meishan pigs showed that ART3 is mostly expressed in muscles enriched in fast twitch fibers^47,48^, which are typically more prone to damage in DMD. ART3 has been reported to be decreased in the serum across multiple dystrophies and myopathies^39^ and in SBMA^45^. It has been recently reported in SBMA only 3 proteins, including RGMA, MSTN, and ART3 were found to have decreased abundances in plasma and were significantly correlated with higher thigh MRI muscle fat fraction akin to what is observed in DMD^45^. Additional support for the relationship to fat deposition has been obtained in Wannanhua pigs show that ART3 is involved in fat deposition in muscle^49^, further strengthening the biological rationale behind the association identified in this study. This is the first study to show ART3 relationship to CS status and clinical function in DMD.

Finally, EHMT2, known as euchromatic histone lysine methyltransferase 2, and DLK1 known as Delta-like 1 homolog as well as Pref-1 (preadipocyte factor 1) showed interesting association. They both declined with age and were associated with disease milestone; EHMT2 was normalized by steroid treatment. EHMT2 was previously associated with renal fibrosis^50^, atrial fibrosis^51^, cardiomyocytes hypertrophy^52^ and high fat diet induced obesity and hepatic insulin resistance^53^, which align with the pathogenic processed ongoing in DMD. The reduction of EHMT2 with age could potentially describe the reduced magnitude of pathological processes as muscle mass is progressively lost. DLK1 plays a multifaceted role in muscle development and regeneration. DLK1 is a transmembrane protein that functions as a regulator of cell growth during development. In adults, its expression is low and restricted to endocrine tissues. It is involved in the differentiation of multiple cell types, including adipocytes and plays an important role in skeletal muscle biology during fetal development and postnatal growth^54^. While the role of DLK1 in adult skeletal muscle regeneration is less clear, upregulated expression of DLK1 has previously been observed in DMD and Becker muscular dystrophy^55^ and reduction of DLK1 in fibroadipogenic progenitors corresponded to increased adipogenic committment^56^. The reduction of the DLK1 levels may be associated with an increased adipogenic signature in DMD.

One limitation of this study was the differences in cohort characteristics due to the participant pool and standards of care. The LUMC samples consisted of participants seen in the clinic, were generally younger in age, and were primarily undergoing a 10 days on, 10 days off CS regimen. No data were available on whether a patient was in the “on” or “off” phase of treatment at the time of sample collection. In contrast, the UF cohort samples were from participants in a natural history research study, were generally older in age, and were primarily on a daily CS regimen. The differences in steroid regimens may have led to smaller effect sizes associated with CS use in the LUMC cohort as well as the earlier occurrence of clinical milestones. The smaller proportion of individuals reaching milestones in the UF cohort may explain the smaller number of probes associated with clinical outcomes and milestones in that cohort. As different regimes coincided with cohort effects, we were unable to study the impact of intermittent CS compared to daily CS. These differences could potentially be studied in prospective research such as in the FOR-DMD clinical trial. Lastly, due to the retrospective nature of this study, we had a number of missing data points. Despite these limitations, due to our relatively large patient population and number of samples, we were able to identify multiple significant serum biomarkers which were validated across both cohorts.

In conclusion, we found proteins associated with upper and/or lower limb outcomes in individuals with DMD across two independent cohorts. Significant consideration should be given to the use of RGMA, DLK1, ANTXR2, EHMT2 and ART3 as potential biomarkers based on the strength of their associations, significance of the findings across scales and milestones and biological plausibility in connection to the disease. A panel that accurately and precisely detects these proteins in serum could enable the connection of short-term changes to disease stabilization and a decreased risk of decline in the mid-to long-term. Furthermore, measuring these proteins in a clinical setting would help predict individual disease trajectories, assess treatment options before milestones, and inform patient inclusion criteria in clinical trials. These findings are of importance for future clinical trial design, paving the way for innovative outcome measures such as serum biomarkers.

## Acknowledgements

This study was funded by Parent Project Muscular Dystrophy through the Protein Mapping Project. Research reported in this publication was also partially supported by Spieren voor Spieren (grant number Svs15) and National Institute Of Neurological Disorders And Stroke of the National Institutes of Health under Award Number # R61NS119639 (Co-I: Spitali), R01 AR056973 (PI: K Vandenborne), and R01 AR065943 (PI: GA Walter). Research reported in this publication was also supported by the CTSI Biorepository at the University of Florida Clinical and Translational Science Institute, which is supported in part by the NIH National Center for Advancing Translational Sciences under award number UL1 TR001427. The authors thank the staff and participants for their contributions.

